# Genomic characterization of therapy-associated polyposis reveals an alkylating mutational signature from prior treatment

**DOI:** 10.64898/2026.02.12.25340205

**Authors:** Yashika Parashar, Zsofia Sztupinszki, Aurel György Prósz, Xiaolu Wang, Pratyusha Bala, Shweta R. Cavale, Chinedu Ukaegbu, Sapna Syngal, Asaf Maoz, Leah Biller, Ramona Lim, Matthew B. Yurgelun, Zoltan Szallasi, Nilay Sethi

**Affiliations:** Division of Molecular and Cellular Oncology, Department of Medical Oncology, Dana-Farber Cancer Institute, Boston, MA, USA; Department of Medicine, Harvard Medical School, Boston, MA, USA; Danish Cancer Institute, Copenhagen, Denmark; Computational Health Informatics Program, Boston Children’s Hospital, Boston, MA; Division of Cancer Genetics and Prevention, Department of Medical Oncology, Dana-Farber Cancer Institute, Boston, MA, USA; Department of Systems Biology, Harvard Medical School, Boston, MA, USA; Division of Gastrointestinal Oncology, Department of Medical Oncology, Dana-Farber Cancer Institute, Boston, MA, USA; Division of Gastroenterology, Hepatology, and Endoscopy, Brigham and Women’s Hospital, and Harvard Medical School; Broad Institute of Massachusetts Institute of Technology (MIT) and Harvard University, Cambridge, MA, USA

## Abstract

Gastrointestinal (GI) polyposis is a major risk factor for colorectal cancer (CRC) and a defining feature of hereditary polyposis syndromes such as familial adenomatous polyposis (FAP). Therapy-associated polyposis (TAP), however, is a rare and incompletely characterized condition that develops decades after treatment for childhood or young adult cancers (CYAC), most often following abdominopelvic radiation or exposure to alkylating agents. As long-term CYAC survival improves, the burden of late GI toxicity, including markedly elevated risks of polyps, CRC, and secondary cancers, continues to rise, yet the molecular features of TAP remain poorly understood.

Here, we present the largest clinicopathological and genomic study of TAP to date, comprising 29 patients diagnosed at a median age of 49 years and a median latency of 29 years after primary cancer therapy. Most patients (78%) had received alkylating agents and exhibited high rates of secondary malignancies.

Histopathology revealed mixed polyp subtypes with a predominance of adenomas. Given these features and the presence of family history in a subset of patients, we investigated the possibility of Hereditary Mixed Polyposis Syndrome (HMPS). Whole-genome sequencing excluded HMPS by demonstrating absence of the canonical 40-kb GREM1 duplication and lack of consistent GREM1 overexpression. Comparative genomic analysis revealed that TAP adenomas exhibit more extensive genome fragmentation and a higher burden of large structural variants than FAP adenomas. Mutational signature profiling identified strong contributions from age-associated signatures (SBS1, SBS5) and a strong, pervasive contribution of the alkylating-agent signature SBS25, even in samples lacking matched normal tissue, whereas platinum-associated SBS31 was minimal. Patient-derived organoids from TAP adenomas showed impaired differentiation, suggesting persistent therapy-induced stem cell dysfunction.

Together, these findings define TAP as a distinct polyposis syndrome marked by heterogeneous histology, long latency, profound structural genomic injury, and chemotherapy-specific mutational scars. This work supports early and tailored GI surveillance for CYAC survivors and provides mechanistic insight into the long-term consequences of cytotoxic therapy on intestinal epithelial homeostasis.

## Manuscript

Gastrointestinal (GI) polyposis is a well-established risk factor for colorectal cancer (CRC) and a hallmark of hereditary syndromes such as familial adenomatous polyposis (FAP), which results from germline APC mutations. Therapy-associated polyposis (TAP) is a rare, uncharacterized form of GI polyposis that emerges decades after treatment for childhood or young adult cancers (CYAC), typically involving abdominopelvic radiation and/or alkylating chemotherapy. Unlike hereditary syndromes, TAP lacks a defined genetic etiology, and its biological underpinnings remain poorly understood.(1)

As long-term survival after CYAC continues to improve, so does the burden of late GI toxicities. Multiple studies now report increased rates of polyps and CRC among CYAC survivors(2), who face nearly an 11-fold increased risk of CRC compared to the general population (3). In one study, 44.4% of CYAC survivors treated with abdominopelvic radiotherapy developed polyps, with 27.8% classified as precancerous.(4) Similarly, the DCOG-LATER cohort found higher adenoma rates in CYAC survivors compared to their siblings.(5) CYAC survivors also face increased risks of other cancers, including thyroid, lung, and breast.(6) Despite these risks, the molecular features of GI polyposis in this population have not been systematically described.

Here, we present a clinicopathological and genomic analysis of 29 patients with TAP, the largest such study to date (**Figure 1A–D**, Supplemental Table 1). The median age at diagnosis of the primary cancer was 16 years (IQR 13, 21). Hodgkin’s lymphoma was the most common primary malignancy, accounting for 65.5% (n=19) of cases (**Figure 1A**). Among those with available treatment history, 78.2% (18 of 23) had received alkylating chemotherapy agents - specifically mechlorethamine, procarbazine, dacarbazine, or cyclophosphamide - during their initial cancer treatment. Abdominal field radiation, either partial or whole, was administered to 36% (9 of 25) of patients who received radiotherapy.

**Figure 1:**
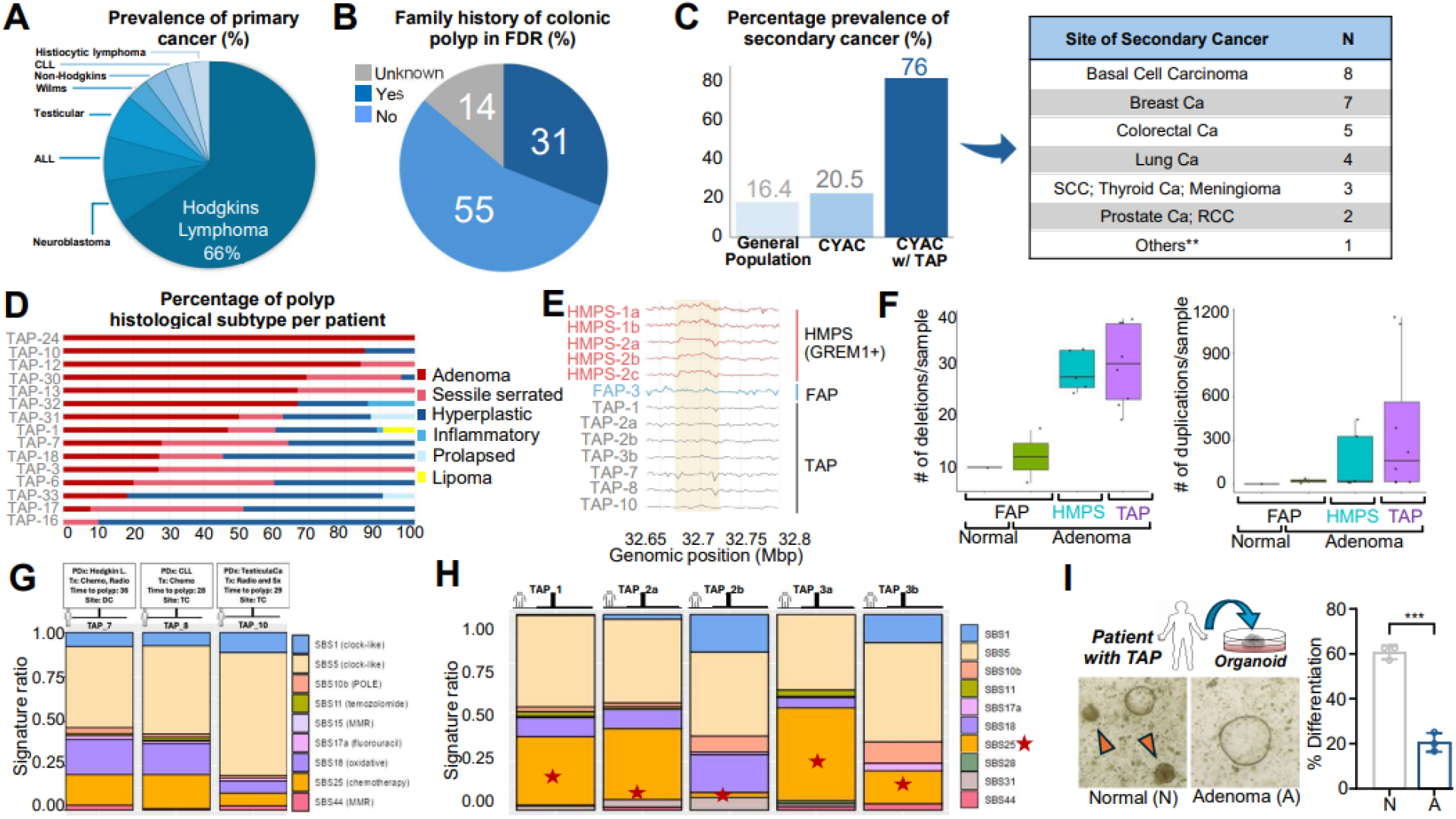
Clinicopathological and genomic analysis of patients with TAP. **A**. Prevalence of primary cancer diagnosis in patients with TAP (n=29). Neuroblastoma (7%); Acute Lymphoblastic Leukemia (7%); Testicular cancer (7%); Wilms (4%); Non-Hodgkins (3%); Chronic Lymphoblastic Leukemia (3%); Histiocytic lymphoma (3%). **B**. Fraction of TAP patients with family history of colonic polyp in first degree relatives (n=29) **C**. Prevalence of secondary cancers in the general population (More than 2 million survivors), childhood and young adult cancer survivors (CYACs) (n = 14,359), and CYACs with TAP (n=29), with distribution of secondary cancer types shown for TAP cases. **Others: Melanoma; Cholangiocarcinoma; AML; DLBCL; Esophageal Ca; Vocal cord SCC; Osteochondroma; Acoustic neuroma; Mandibular schwannoma **D**. Distribution of colonic polyp histology observed in TAP patients (n=15). **E**. Coverage tracks across GREM1 genomic region comparing HMPSS samples with confirmed GREM1 duplication (n=4, red), FAP samples (n=1) and TAP samples (n=8). Each horizontal trace shows per-bin coverage for one sample (bins = 100 bp), normalized by the per-sample median and plotted as log2 ratio. The shaded rectangle marks the region previously reported to harbor duplications affecting GREM1. **F**. Comparison of chromosomal alterations between normal tissue and adenomas in FAP and TAP patients. Boxplots display the number of deletions (left) and duplications (right) per sample, with TAP adenomas showing increased copy number variation compared to normal tissue and FAP adenomas. **G**. Mutational signature analysis of TAP samples with paired germline controls, showing the relative contribution of single base substitution (SBS) signatures. **H**. Mutational signature analysis of TAP samples without paired germline controls, showing the relative contribution of SBS signatures. Somatic mutations were filtered using public databases (see Supplementary Methods). **I**. Light microscopy of paired normal and adenoma organoids from a TAP patient (left); orange arrows indicate differentiated organoids. Quantification of differentiated organoids in normal versus adenoma cultures (right). PDx= primary cancer, Tx= treatment received for primary cancer, Chemo=chemotherapy, Radio=radiotherapy, Sx=surgery, Time to polyp= time between treatment of primary cancer and detection of first polyp (in years), Site: TC: Transverse colon, DC: Descending colon, Hodgkin L=Hodgkin’s lymphoma, CLL=Chronic Lymphocytic Leukemia.

9 patients (31%) had family history of colonic polyp in first degree relatives, including 2 patients with additional family history of polyps in second degree relatives (**Figure 1B**). Notably, there is a markedly elevated frequency of secondary cancers among patients with TAP, more than the general and non-TAP CYAC populations. Basal cell carcinoma was diagnosed in 8 patients (27.5%) while 7 females (50%) reported breast cancer. Colorectal cancer (n=5) and lung cancer (n=4) were commonly reported secondar cancers in the cohort (**Figure 1C**).

TAP was diagnosed at a median age of 49 years (IQR 44, 55), substantially earlier than the typical age of polyposis diagnosis in the general population.(7) The median latency between primary cancer treatment and polyposis detection was 29 years (IQR 26, 36). On average, patients underwent 5.4 colonoscopies, highlighting the considerable burden of long-term GI surveillance. Histopathologic evaluation revealed a predominance of mixed polyp subtypes, most frequently adenomatous in nature **(Figure 1D)**.

Given the mixed histology of colonic polyps, the presence of secondary cancers, and a family history of colonic polyps in 9 patients, we considered the possibility of Hereditary Mixed Polyposis Syndrome (HMPS), which is driven by a germline duplication upstream of *GREM1* that leads to greater GREM1 expression.(8) To test this hypothesis, we performed whole genome sequencing (WGS) of adenomas from patients with HMPS (n=4 from 2 patients, positive control), familial adenomatous polyposis (FAP, n=2, negative control), and TAP (n=8) with sufficient coverage for structural variant analysis). All four HPMS samples showed the 40-kb *GREM1* duplication. (**Figure 1D**). While *GREM1* duplications were found in each HMPS case, it was not observed in TAP cases. (**Figure 1E, Supplementary Table 1**.). Calls were supported by normalized read-depth graphs, CNVnator, and manual review in IGV. We also assessed GREM1 expression in representative adenomas from each group by immunohistochemistry and observed heterogeneous expression without evidence of consistent overexpression in TAP (Supplemental Figure 1).

After excluding low coverage TAP samples, detailed genomic analyses were performed on four groups: FAP normal mucosa (n = 1), FAP polyps (n = 2), HMPS polyps (n=5) and TAP polyps (n = 8). TAP adenomas harbored a greater number of large deletions and insertions (>1 kb) and exhibited significantly more genome fragmentation than FAP adenomas. (**Figure 1F**). To assess whether TAP adenomas bear mutational scars attributable to prior CYAC therapy, we conducted mutational signature analysis. All samples showed consistent contributions from age-related signatures SBS1 and SBS5, which together accounted for 45–70% of mutations per sample. Notably, SBS5 was consistently enriched across all TAP samples, suggesting an elevated age-associated or possibly treatment-influenced background mutational burden. Importantly, SBS25, a signature linked to prior chemotherapy treatment, was robustly detected in TAP adenomas, including in samples lacking matched normal tissue (**Figure 1G-H**). The platinum-associated SBS31 signature remained minimal in all cases.

SBS25 emerged as a particularly notable finding, as it is not typically observed in colorectal cancer or premalignant lesions. While initially identified in Hodgkin lymphoma cell lines with an unclear origin, recent whole-genome sequencing studies of Hodgkin and Reed–Sternberg cells have reinforced its association with prior therapy. Specifically, SBS25 has been detected exclusively in tumors from individuals treated with procarbazine or dacarbazine, which are both methyl-hydrazine-based alkylating agents.(9) Mechanistically, SBS25 is characterized by T>A transversions, a mutational pattern consistent with DNA mispairing lesions induced by methyl-hydrazine alkylation.

To further explore functional consequences of TAP, we generated paired normal and adenoma organoids from a TAP patient. The TAP adenoma organoid exhibited impaired differentiation relative to its matched normal counterpart, phenocopying the differentiation defects observed in FAP adenoma organoids (**Figure 1I**).(10) This system offers a tractable model for future mechanistic interrogation of TAP biology.

Together, these findings position TAP as a complex clinical and molecular entity within the spectrum of polyposis syndromes, with a multifaceted etiology attributable to prior chemotherapy, radiation exposure, or unexposed germline predisposing alterations. The syndrome is defined by its mixed polyp histology and manifestation of polyposis following CYAC therapy. Our genomic analysis showed the presence of chemotherapy-associated mutational signatures in most TAP adenomas, although further mechanistic work is required to clarify causality. Patient-derived organoid models, as demonstrated here, provide a valuable platform to dissect the genetic and epigenetic underpinnings of TAP. Given that most TAP cases do not harbor known germline alterations linked to polyposis, early surveillance for gastrointestinal neoplasia and tailored screening strategies should become an integral component of survivorship care in CYAC patients.

### Study limitations

These conclusions must be interpreted in the context of certain limitations, including small sample size, lack of matched normal samples for many TAP cases, and variability in sequencing depth, which may affect the accuracy of CNV calling.

## Supporting information

Supplemental Section

## Data Availability

All data produced in the present study are available upon reasonable request to the authors

## Notes

**Conflicts of Interest:** N.S.S. receives research funding from Novartis, is a consultant for Dewpoint Therapeutics, and is on the scientific advisory board for Astrin Biosciences

### Competing Interest Statement

N.S.S. receives research funding from Novartis, is a consultant for Dewpoint Therapeutics, and is on the scientific advisory board for Astrin Biosciences

### Funding Statement

This study did not receive any funding

### Author Declarations

Ethics committee/IRB of Dana Farber Cancer Institute and Harvard Medical School gave ethical approval for this work

