## Supplemental Section for "Genomic characterization of therapy-associated polyposis reveals an alkylating mutational signature from prior treatment"

### Supplemental Material

### Supplemental Figures

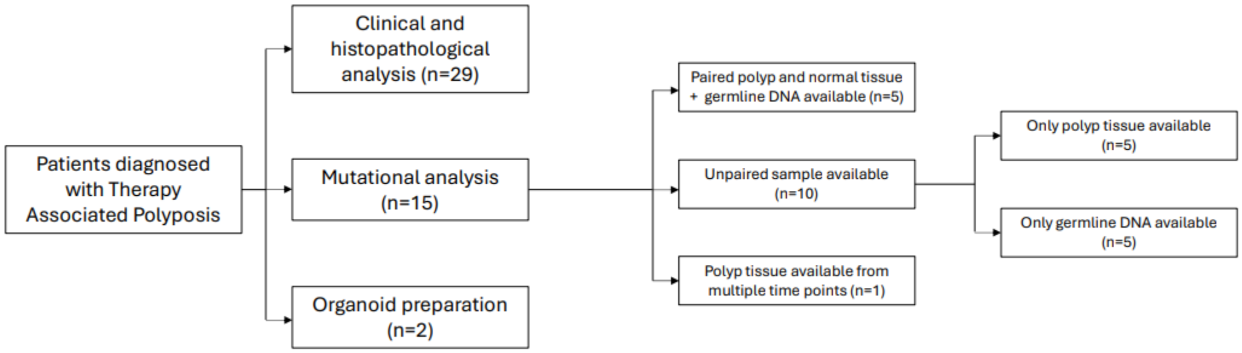


**Supplemental Figure 1.** Consort diagram of the study population included in the study.

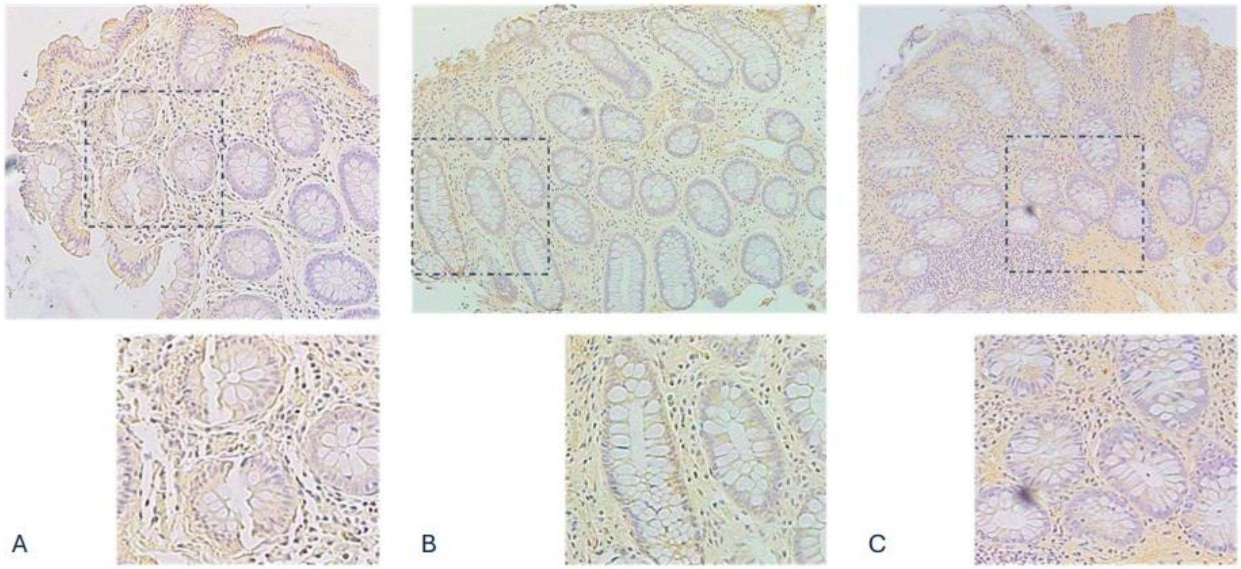


**Supplemental Figure 2.** Representative images of anti-GREM1 immunohistochemistry of colonic polyps from (A) a patient with therapy associated polyposis, B) a patient with known GREM1 duplication, C) a patient with familial adenomatous polyposis.

**Supplemental Methods:**

Patients with Therapy Associated Polyposis enrolled at Dana Farber Cancer Institute, Brigham and Women's Hospital and Massachusetts General Hospital between 2014- 2023 were identified and included in the study. Polyposis was defined as the lifetime incidence of 10 or more gastrointestinal polyps, and individuals diagnosed with CYAC at age 30 or between ages 31–45, provided their first gastrointestinal polyp was identified ten years after initial CYAC treatment, were included in the study. Patients with personal or family diagnoses of pathogenic germline variants in genes linked to inherited colorectal cancer susceptibility were excluded. The Institutional Review Board at DFCI approved the study and informed written consent was obtained from all patients. Baseline characteristics of the population including age, sex, race, ethnicity, mean age of primary cancer, type of primary cancer, treatment received for primary cancer, including site of radiotherapy and type of chemotherapy received, mean age of development of first polyp, and detailed histology of polyps observed, were annotated retrospectively from electronic health records. Additionally, history of secondary cancers, other systemic side effects of exposure to radiotherapy and chemotherapy and history of cancers in first degree relatives and second-degree relatives were recorded. We investigated the overexpression of GREM1 protein using Immunohistochemistry with Anti GREM1 antibodies on formalin fixed paraffin embedded (FFPE) tissue slides obtained for 2 (28.6%) out 10 patients (34.5%) with family history of colonic polyposis in first degree relatives among the 29 patients with TAP.

To identify genomic alterations associated with TAP, we conducted Whole Genome Sequencing on polyp samples for 15 patients (51.7%) with TAP. Samples for DNA isolation were collected retrospectively from archived frozen blocks containing polyp tissue for 9 patients, including 2 patients with polyp tissue from more than 1 site. Fresh polyp samples along with normal tissue were collected for 1 patient during colonoscopy. DNA isolation from FFPE samples was performed using the QIAamp DNA FFPE tissue kit while QIAamp DNA Micro Kit was used to extract DNA from the fresh polyp sample.

Mutect2 was used to detect point mutations and indels in every sample (GATK 4.5.0.0), after aligning to the hg38 (GCA_000001405.15_GRCh38) reference genome. Only samples with a median coverage of at least 10 were evaluated. For samples without a paired normal sample (both WGS and WES cases), only variants with an allele frequency of less than 25% were regarded as possible somatic variants. Furthermore, these samples were further filtered for variants which had more than 5 tumor ALT read count and this list was refined more by retaining those mutations that are not present in gnomAD (V2.1) or KAVIAR (version 160204-Public) with an allele frequency greater than 0.01. Additionally, clustered mutations were also removed. To filter out the remaining variants caused by technical errors, the intersection of mutations between all the samples was also not included in the final call set.

Single base substitution (Cosmic V3 SBS, filtered for the colorectal cancer signatures, based on: https://www.nature.com/articles/s41586-020-1943-3) signatures were extracted using the deconstructSigs and MutationalPatterns R packages (version 1.9.0, and 3.16.0 respectively), the short indel and doublet base substitution (ID and DBS) signatures were extracted using the ICAMS R package (version 2.3.12). Treatment-associated signatures have been also extracted alongside the colorectal cancer signatures, and included: SBS11, SBS25, SBS18, SBS36, SBS87, SBS31) These identify the linear combination of pre-defined signatures that best reconstructs a tumor's mutational profile.

In case of whole-genome samples copy number, changes were evaluated using depth- and segmentation-based callers and then merged into a single high-confidence catalogue. CNVnator (v0.4.1; bin size = 100 bp) generated read-depth segments with associated quality metrics, including the fraction of zero-mapping reads (q0) and CNVkit (v0.9.10) output $log_2$ copy-ratio segments for each sample. Both call sets were imported into R 4.3 (GenomicRanges, data.table) and low-confidence regions and segments shorter than 1 kb were excluded. Because CNVkit exhibits finer break-point resolution, its segments were first symmetrically padded by ±20 bp to allow for caller-specific rounding differences, then intersecting regions of the same CNV type (gain or loss) were identified. To identify and exclude GREM1 duplications CNVnator and delly was used and the reads were manually inspected using IGV. For visualization the normalized log2 copy-ratio profiles coverage using 3kb bins and default CNVkit parameters were plotted.

Tissue specimen from an unidentifiable patient with TAP was collected post endoscopy under approval (14-408) by the Internal Review Board of the Dana Farber Cancer Institute, Boston, Massachusetts, USA. For generation of organoids from the patient-derived and mouse-derived tissues, the tissues were treated with EDTA and then resuspended in 30–50 μl of Matrigel (BD Bioscience) and plated in 24-well plates. WNT/R-spondin/Noggin (WRN) containing DMEM/F12 with HEPES (Sigma-Aldrich) containing 20% FBS, 1% penicillin/streptomycin and 50 ng/ml recombinant mouse EGF (Life Technologies) was used for culturing colon organoids. For the first 2–3 days after seeding, the media was also supplemented with 10 mM ROCK inhibitor Y-27632 (Sigma Aldrich) and 10 mM SB431542 (Sigma Aldrich), an inhibitor for the transforming growth factor (TGF)-β type I receptor to avoid anoikis. For passage, colon organoids were dispersed by trypsin-EDTA and transferred to fresh Matrigel. Passage was performed every 3-4 days with a 1:3–1:5 split ratio. For human colon organoid culture, the previous media was supplemented with antibiotics 100 μg/ml Primocin (Invivogen), 100 μg/ml Normocin (Invivogen); serum-free supplements 1× B27 (Thermo Fisher (Gibco)), 1X N2 (Thermo Fisher (Gibco)); chemical supplements 10 mM Nicotinamide (Sigma), 500mM N-acetylcysteine (Sigma), hormone 50 mM [Leu15]-Gastrin (Sigma), growth factor 100 μg/ml FGF10 (recombinant human) (Thermo Fisher) and 500nM A-83-01 (Sigma), which is an inhibitor of the TGF-β Receptors ALK4, 5, and 7.
